# Evolutionary divergence and expression of differential HLA alleles between donor and recipient influence acute GVHD onset after allogenetic HSCT

**DOI:** 10.64898/2025.12.10.25342012

**Authors:** Yingjie Chen, Wenqi Zhan, Ying Zhang, Hua Gao, Yue Han, Jun Qiao, Zihang Li, Yundie Liu, Yuanyuan Li, Shuhan Yang, Yaxin Guo, Xinyu Wang, Fei Li, Xiaoning Wang, Pengcheng He, Peng Gao

**Affiliations:** Department of Hematology, The First Affiliated Hospital of Xi’an Jiaotong University, Xi’an, China; Department of Hematology, The First Affiliated Hospital of Nanchang University, Nanchang, China; School of Computer Science and Technology, Zhejiang University of Water Resources and Electric Power, Hangzhou, China; Department of Pharmacology, School of Medicine, Southern University of Science and Technology, Shenzhen, China; Center for Mitochondrial Biology and Medicine, The Key Laboratory of Biomedical Information Engineering of Ministry of Education, School of Life Science and Technology, Xi’an Jiaotong University, Xi’an, China; Genome Institute, The First Affiliated Hospital of Xi’an Jiaotong University, Xi’an, China

**Keywords:** HLA evolutionary divergence, expression, hematopoietic stem cell transplantation, acute graft-versus-host disease, dendritic cell

## Abstract

Acute graft-versus-host disease (aGVHD) remains a major complication after hematopoietic stem cell transplantation (HSCT), especially given haploidentical HSCT is now China’s primary method, yet this context lacks reliable predictors. aGVHD initiation, involving donor T cell activation by recipient conventional dendritic cells (cDCs), depends on donor-recipient HLA disparity and its expression. We therefore developed the donor and recipient-specific HLA evolutionary divergence (DRs_HED) algorithm to quantify relevant HLA differences, then integrated HLA expression in cDC to evaluate the combined predictive value. While DRs_HED correlated with aGVHD occurrence, links to organ severity varied: consistent for skin but not gastrointestinal aGVHD. Only cDCs expression of HLA-DRB1 correlated with presentation of associated minor histocompatibility antigens (miHAs). Integrating expression with DRs_HED improved prediction, raising the model’s AUC by about ∼10% to 0.607 ± 0.012 versus DRs_HED alone. Our findings show precise quantification of HLA disparity and expression improves aGVHD risk prediction, aiding future multi-factorial models.

## INTRODUCTION

Allogeneic hematopoietic stem cell transplantation (aHSCT) is a curative therapy for various hematological malignancies, bone marrow failure syndromes, and immunodeficiencies^1,2^. However, its application still faces numerous challenges, notably acute graft-versus-host disease (aGVHD).^3,4^. aGVHD typically develops within 100 days post-transplant, driven by donor T cells that recognize and attack recipient tissues. The skin, gastrointestinal tract, and liver are the primary targets, with severe cases leading to multi-organ failure or death^3,5,6^. Despite advancements in prophylaxis and treatment, aGVHD remains a major determinant of post-transplant survival and quality of life^7–10^.

The main risk factors for aGVHD include donor-recipient HLA mismatch, age, sex disparities, and conditioning intensity, with HLA compatibility being the most critical^11,12^. Although aGVHD risk exhibits substantial correlation with the number of mismatched HLA loci^13,14^, clinical outcomes often vary significantly even among HLA-matched transplants^15^, partly due to differences in minor histocompatibility antigens (miHAs) presented by HLA molecules^16–18^ Nevertheless, despite growing evidence of their critical role in regulating immune activity^19–21^, the degree of difference between mismatched HLA alleles and their expression levels have been overlooked in donor selection and aGVHD prediction.

Recently, HLA evolutionary divergence (HED) has emerged as a novel metric to quantify functional differences between HLA alleles based on Grantham distance which measures physicochemical variation in antigen-binding domains^22^. Across diverse immunological contexts, including cancer immunotherapy, solid organ transplantation, and infectious diseases, HED has been shown to correlate with immune response strength and clinical outcomes^21,23–25^. In aHSCT, recipient HED or the sum of donor-recipient pairwise HED values has been associated with graft-versus-leukemia effects, relapse, and survival, though results are inconsistent., possibly because current algorithms fail to accurately capture donor-recipient specific HLA differences in the context of aGVHD^26–30^.

In addition to genetic divergence, the role of HLA expression levels in immune regulation has received increasing attention^19,31^. High expression of mismatched *HLA-C* alleles in recipient T cells is associated with severe aGVHD and mortality, suggesting that elevated HLA-C expression promotes donor T cell activation and triggers a strong response^32,33^. Similar effects have been observed at mismatched *HLA-DPB1* loci^34,35^. Nevertheless, the role of HLA allele expression in conventional dendritic cells (cDCs), the key antigen-presenting cells initiating aGVHD, remains unclear.

Here, we established the DRs_HED algorithm to precisely calculate donor-recipient HLA differences under the immunological context of aGVHD. We further integrate HLA allele expression levels in cDCs to investigate the individual and combined effects of HLA divergence and expression on aGVHD outcomes, aiming to provide a basis for more accurate donor selection and risk stratification strategies in the future.

## RESULTS

### Patient baseline characteristics

This study cohort comprised 774 patients who underwent aHSCT between 2013 and 2024, with 503 cases from the First Affiliated Hospital of Xi’an Jiaotong University and 271 from the First Affiliated Hospital of Nanchang University. The median patient age was 34 years (range, 2-69 years), with 58.7% males and 41.3% females. The primary disease indications were acute myeloid leukemia (AML, n = 314), acute lymphoblastic leukemia (ALL, n = 208), myelodysplastic syndrome (MDS, n = 101), and aplastic anemia (AA, n = 119). Peripheral blood was the predominant stem cell source (64.2%), and haploidentical transplants accounted for the majority (70.2%). Most patients received myeloablative conditioning (MAC, 84.6%). Before transplantation, 553 patients were in complete remission, and 448 were MRD-negative. Donor-recipient CMV serostatus was predominantly double-positive (756 pairs, 97.7%). Male donors comprised 68.1% (n = 527), with male-to-male donor-recipient pairs being the most common combination (39.4%, 305 pairs). Detailed baseline characteristics are summarized in Table 1.

**Table 1.** Baseline characteristics of aHSCT recipients and donors.

| Variable | Number/Median | %/Range |
| --- | --- | --- |
| All patients | 774 | 100 |
| Center |  |  |
| The First Affiliated Hospital of Xi'an Jiaotong University | 503 | 65.0 |
| The First Affiliated Hospital of Nanchang University | 271 | 35.0 |
| Gender |  |  |
| Female | 320 | 41.3 |
| male | 454 | 58.7 |
| Recipient age (median and range) | 34 | 2-69 |
| Donor age (median and range) | 34 | 4-63 |
| Diagnosis |  |  |
| AML | 314 | 40.6 |
| ALL | 208 | 26.9 |
| MDS | 101 | 13.0 |
| AA | 119 | 15.4 |
| CML | 9 | 1.2 |
| NHL | 9 | 1.2 |
| HLH | 3 | 0.4 |
| MAL | 3 | 0.4 |
| MS | 2 | 0.3 |
| ANKL | 1 | 0.1 |
| APL | 1 | 0.1 |
| CMML | 1 | 0.1 |
| DC | 1 | 0.1 |
| HL | 1 | 0.1 |
| LBL | 1 | 0.1 |
| Graft source |  |  |
| PB | 497 | 64.2 |
| CB | 1 | 0.1 |
| BM | 2 | 0.2 |
| CB+PB | 68 | 8.8 |
| BM+PB | 180 | 23.3 |
| CB+PB+BM | 26 | 3.4 |
| HLA match |  |  |
| Identical sibling | 197 | 25.5 |
| Haplo-identical | 543 | 70.2 |
| Matched Unrelated | 29 | 3.7 |
| Mismatched Unrelated | 5 | 0.6 |
| Conditioning |  |  |
| MAC | 655 | 84.6 |
| RIC | 95 | 12.3 |
| NMA | 19 | 2.5 |
| NA | 5 | 0.6 |
| Remission at aHSCT |  |  |
| CR | 553 | 71.4 |
| PR | 20 | 2.6 |
| NR | 68 | 8.8 |
| NA | 133 | 17.2 |
| Pre-HSCT MRD |  |  |
| Positive | 62 | 8.0 |
| Negative | 448 | 57.9 |
| NA | 264 | 34.1 |
| Donor/recipient CMV match |  |  |
| Negative/positive | 12 | 1.6 |
| Positive/negative | 6 | 0.8 |
| Positive/positive | 756 | 97.6 |
| Donor/recipient gender |  |  |
| Female/male | 136 | 17.6 |
| Others | 638 | 82.4 |

**Abbreviations:** AA: aplastic anemia; ALL: acute lymphoblastic leukemia; AML: acute myeloid leukemia; ANKL: aggressive natural killer cell leukemia; APL: acute promyelocytic leukemia; BM: bone marrow; CB: cord blood; CML: chronic myeloid leukemia; CMML: chronic myelomonocytic leukemia; CR: complete remission; DC: dyskeratosis congenita; HL: Hodgkin lymphoma; LBL: lymphoblastic lymphoma; MAC: myeloablative conditioning; MAL: mixed lineage leukemia; MDS: myelodysplastic syndromes; MRD: minimal residual disease; MS: myeloid sarcoma; NA: not available; NHL: non-Hodgkin lymphoma; NMA:
non-myeloablative conditioning; NR: no remission; PB: peripheral blood; PR: partial remission; RIC: reduced intensity conditioning.

### DRs_HED better reflects donor-recipient HLA allele differences affecting aGVHD occurrence

The antigen-binding domains of HLA alleles feature distinct amino acid sequences that specify unique peptide-binding repertoires. Therefore, the HED between an individual’s two HLA alleles reflects that individual’s antigen-presenting capacity. Although initially applied to assess cancer patients’ responsiveness to immune checkpoint inhibitor (ICI) therapy^21^, this metric requires adjustment for prediction of aGVHD which largely arises from HLA mismatches between donor and recipient. Therefore calculating HED based solely on recipient’s HLA alleles (hereinafter referred to as R_HED) is inadequate in this context. Appling of R_HED to our cohort also showed no significant correlation between aGVHD incidence and single gene or averaged HED values across six HLA loci (**Figure S1A**). To address this limitation, donor-recipient HED (DR_HED) was introduced, defined as the cumulative divergence across all donor-recipient allele pairs^28^. In our cohort, DR_HED of HLA class I loci showed a significant correlation with aGVHD occurrence (**Figure S1B**). Cumulative incidence analysis further revealed significant differences between high- and low-HED groups specifically at class I loci under both methods (**Figure S1C&D**). At the individual gene level, significant associations with aGVHD were confined to *HLA-B* and *HLA-DPB1* (**Figure S2**).

With the widespread adoption of the “Beijing protocol,” haploidentical aHSCT has become the primary transplantation strategy in China. In this setting, each HLA locus typically carries at least one allele shared between the donor and recipient (referred to as “shared HLA”), while the nonshared allele present only in recipient is designated “recipient-specific HLA”. Mechanistically, donor-derived alloreactive T cells are activated by recipient-specific HLA-peptide complexes presented on the surface of recipient antigen-presenting cells (APCs) (**Figure 1A**). Therefore, only the divergence between donor alleles and the recipient-specific HLA accurately reflects the immunological mismatch driving aGVHD. By contrast, the indiscriminate calculation of the sum of all pairwise divergences in DR_HED introduces redundancy in quantifying donor-recipient HLA disparity, potentially compromising its accuracy as a measure of immunogenic differences^28^.

**Figure 1.**
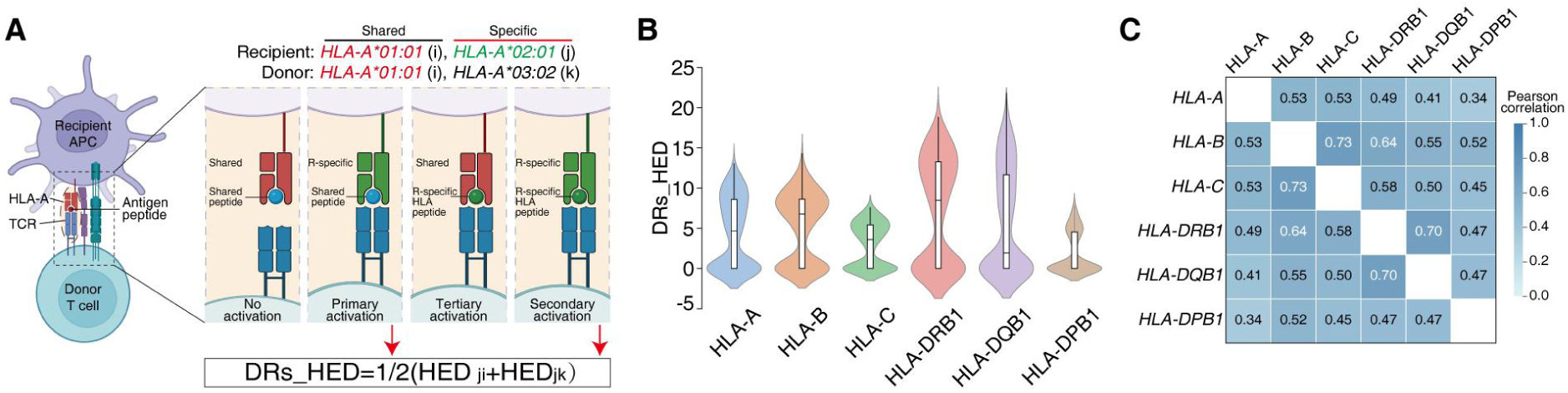
Scheme and descriptive statistics of DRs_HED. (**A**) Schematic illustration of the DRs_HED calculation principle. (**B**) Violin plots showing the distribution of DRs_HED across the 6 HLA genes. (**C**) Correlation matrix of DRs_HED among the 6 HLA genes. APC, antigen-presenting cell; TCR, T cell Receptor.

We therefore proposed a novel metric termed donor and recipient-specific HED (DRs_HED), which specifically quantifies the evolutionary divergence between the recipient-specific HLA allele and both alleles at each locus. For instance, at the *HLA-A* locus, let allele “i” represent the shared allele, “j” the recipient-specific allele, and “k” the donor-specific allele (Figure 1A). The DRs_HED is calculated using the equation (1) as follows:

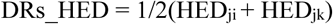

That is, DRs_HED represents the average HED between the recipient-specific HLA gene and the two HLA alleles of the donor at each gene locus. DRs_HED equals zero in their of the following scenarios: when the recipient’s two alleles at a given locus are identical (i=j), or when the donor and recipient are fully matched at the locus (j=k). Under these conditions, donor T cells are not activated by recipient HLA-peptide complexes.

DRs_HED exhibited distinct locus-specific distribution patterns and inter-locus correlations. The distributions of DRs_HED values varied across the six HLA genes: *HLA-C* and *-DPB1* showed relatively narrow distributions, *HLA-DRB1* and *-DQB1* showed the broadest distributions, and *HLA-A* and *-B* fell in between (**Figure 1B**). Correlation analysis revealed moderate inter-locus correlations (r = 0.34-0.73) (**Figure 1C**), with *HLA-B* and *-C* showing the highest correlation (r = 0.73) and *HLA-DRB1* and *-DQB1* also highly correlated (r = 0.70). In contrast, *HLA-DPB1* exhibited relatively weak correlations with other loci (r = 0.34-0.47).

### DRs_HED affects the occurrence and severity of aGVHD

Disparities in HLA alleles between donor and recipient are well-established risk factors for GVHD^14,36^. Therefore, DRs_HED is expected to reflect this correlation more accurately than R_HED and DR_HED. Given the observed inter-locus correlations of DRs_HED, we incorporated each of the six HLA loci individually, along with their overall mean DRs_HED, into regression analyses.

Binary logistic regression revealed that DRs_HED at all three class I loci (*HLA-A*, *-B*, and *-C*) and their mean values were significantly associated with aGVHD incidence (**Figure 2A**). Among class II genes, only *HLA-DRB1* and *HLA-DQB1* reached significance. The six-gene average DRs_HED was also significantly associated with aGVHD risk. Ordinal logistic regression analyzin<u>g</u> aGVHD severity (grades I–IV) yielded largely consistent results, except that *HLA-DPB1* and mean class II DRs_HED were not significant associated. These results indicate that higher cumulative evolutionary divergence correlates with more severe aGVHD (**Figure 2B**). Moreover, using the median single-gene or the average DRs_HED as a cutoff to stratify patients, the high-DRs_HED group consistently exhibited significantly higher aGVHD incidence (**Figure 2C**, **Figure S3**). Additionally, we analyzed relationships between DRs_HED and post-transplant relapse and survival. In contrast to some previous reports, whether individually or on average, DRs_HED generally showed a negative correlation with relapse (**Figure S4A**), but was largely not associated with overall survival and relapse-free survival (**Figure S4B&C**).

**Figure 2.**
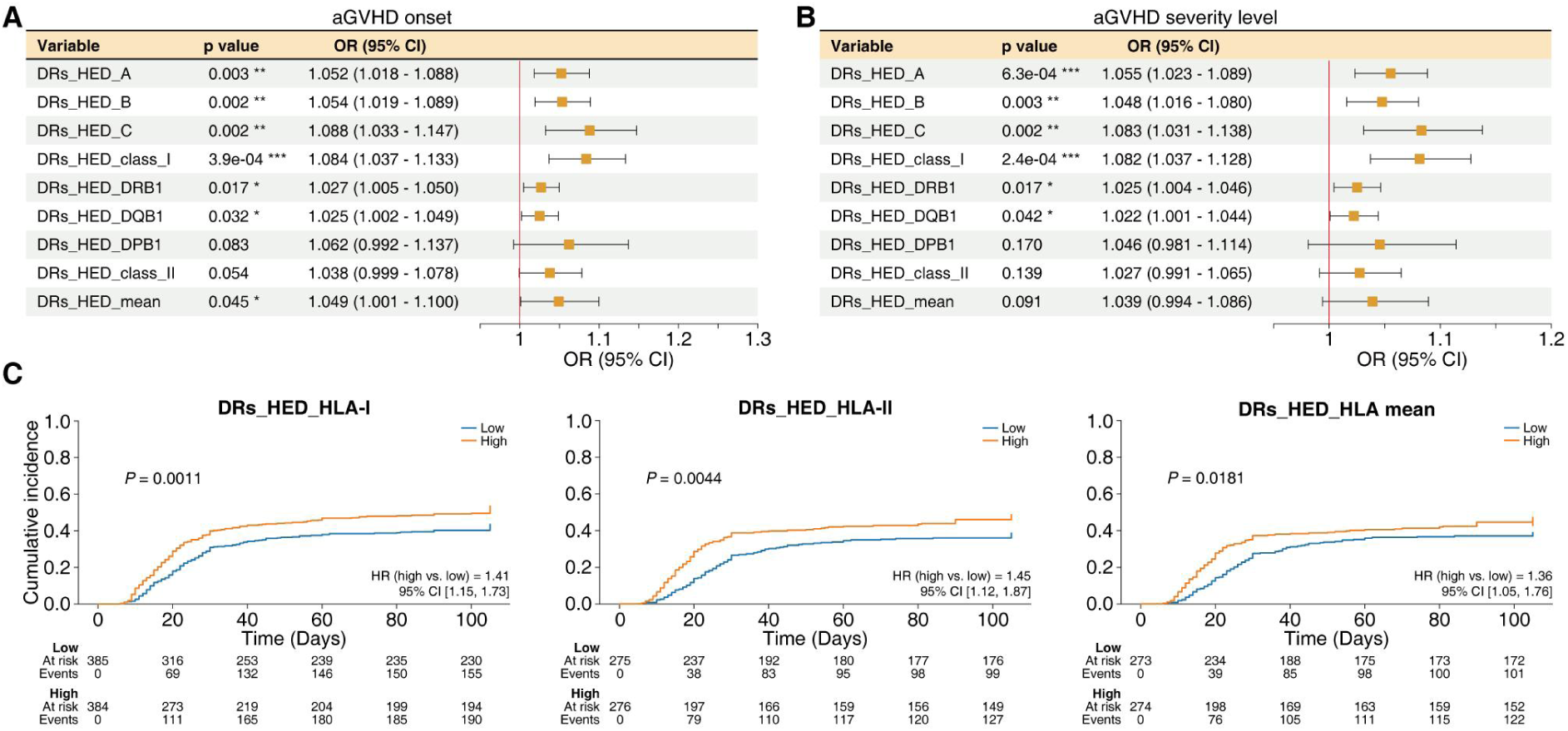
Correlation analysis between DRs_HED and aGVHD. (**A**) Binary logistic regression assessing the association of individual or average DRs_HED of 6 HLA genes with the aGVHD onset. (**B**) Ordinal logistic regression evaluating the relationship between individual or average DRs_HED of 6 HLA genes with aGVHD severity. (**C**) Comparison of cumulative aGVHD incidence between patient groups stratified by the average DRs_HED of HLA-I, HLA-II, and all 6 HLA genes.\**P* < 0.05; \*\**P* < 0.01; \*\*\**P* < 0.001.

In summary, higher DRs_HED levels, whether assessed per gene, by HLA class, or overall, were generally significantly positively correlated with increased aGVHD risk and severity, negatively correlated with relapse, but not associated with survival.

### The correlation between DRs_HED and aGVHD exhibits organ-specific differences

To further investigate whether the association between DRs_HED and aGVHD is universal at the organ level, we analyzed the three principal target organs: skin, gastrointestinal tract, and liver. Strikingly, DRs_HED across all HLA loci, whether considered individually or on average, showed significant association with both the incidence and severity of skin aGVHD, with *HLA-C* exhibiting the strongest effect (OR = 1.106, *P* = 4.2×10^-4^) (**Figure 3A**).

**Figure 3.**
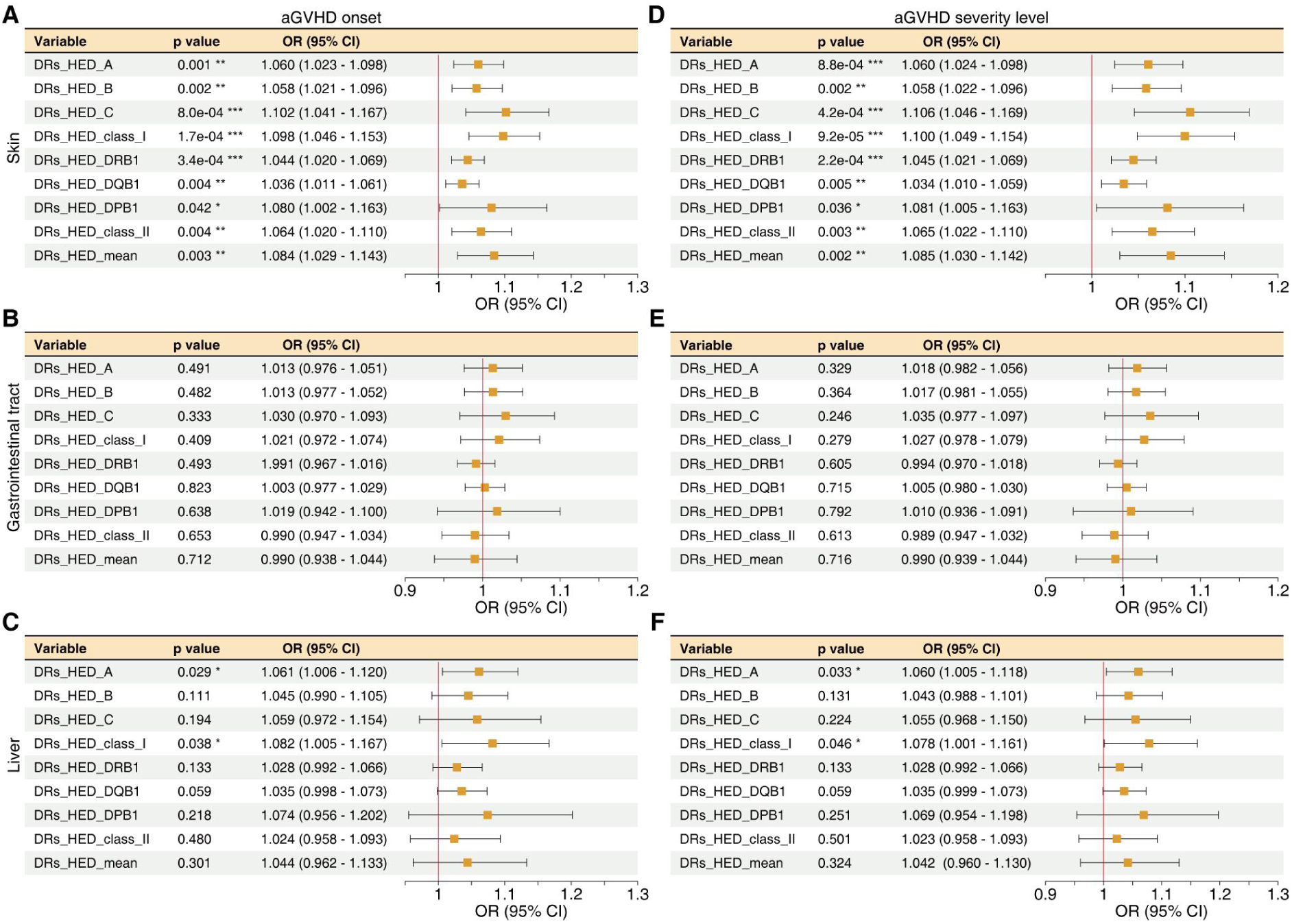
Correlation between DRs_HED and aGVHD in major target organs. (**A-C**) Binary logistic regression assessing association of HLA gene DRs_HED with aGVHD onset in three organs: skin (A), gastrointestinal tract (B), and liver (C). (**D-F**) Ordinal logistic regression assessing the association of HLA gene DRs_HED with aGVHD severity in three organs: skin (D), gastrointestinal tract (E), and liver (F). \**P* < 0.05; \*\**P* < 0.01; \*\*\**P* < 0.001.

In contrast, correlations with gastrointestinal or hepatic aGVHD were markedly weaker and largely non-significant. No HLA locus was significantly associated with gastrointestinal aGVHD (**Figure 3B**). For liver aGVHD, only *HLA-A* DRs_HED displayed a significant association with both occurrence and severity (OR = 1.061, *P* = 0.029) (**Figure 3C**), while the mean class I DRs_HED showed a marginal correlation (OR = 1.082, *P* = 0.038).

In conclusion, the association of HLA gene DRs_HED with aGVHD is highly heterogeneous across major target organs, being strongest for skin aGVHD, weaker for liver aGVHD, and absent for gastrointestinal aGVHD.

### The role of miHAs in aGVHD correlates with the expression level of HLA-DRB1

Although HLA disparity is the principal driver of aGVHD, its relatively high incidence even in fully HLA-matched HSCTs implies an important role for miHAs^3,6^. Theoretically, the impact of miHAs depends on both the extent of donor-recipient disparity and the expression levels of the HLA molecules presenting them. In our study, we can’t quantify the miHA mismatches for limited available information, but we assessed the contribution of HLA allele expression to miHA-mediated aGVHD remains underexplored.

Among residual host APCs post-HSCT, cDCs are the dominant activators of donor T cells^37,38^. As a surrogate for tissue cDCs, we obtained quantitative expression levels of HLA alleles at the single-cell level from peripheral blood cDCs in a public database (see Methods section). Taking the *HLA-C* gene as an example, different alleles showed substantial variation in expression, with certain alleles such as *HLA-C*07:01* also showing considerable individual variability (**Figure 4A**). Due to limited coverage in the dataset with only 4 recipients having all HLA genotypes, subsequent regression analyses were conducted at the single HLA gene level.

**Figure 4.**
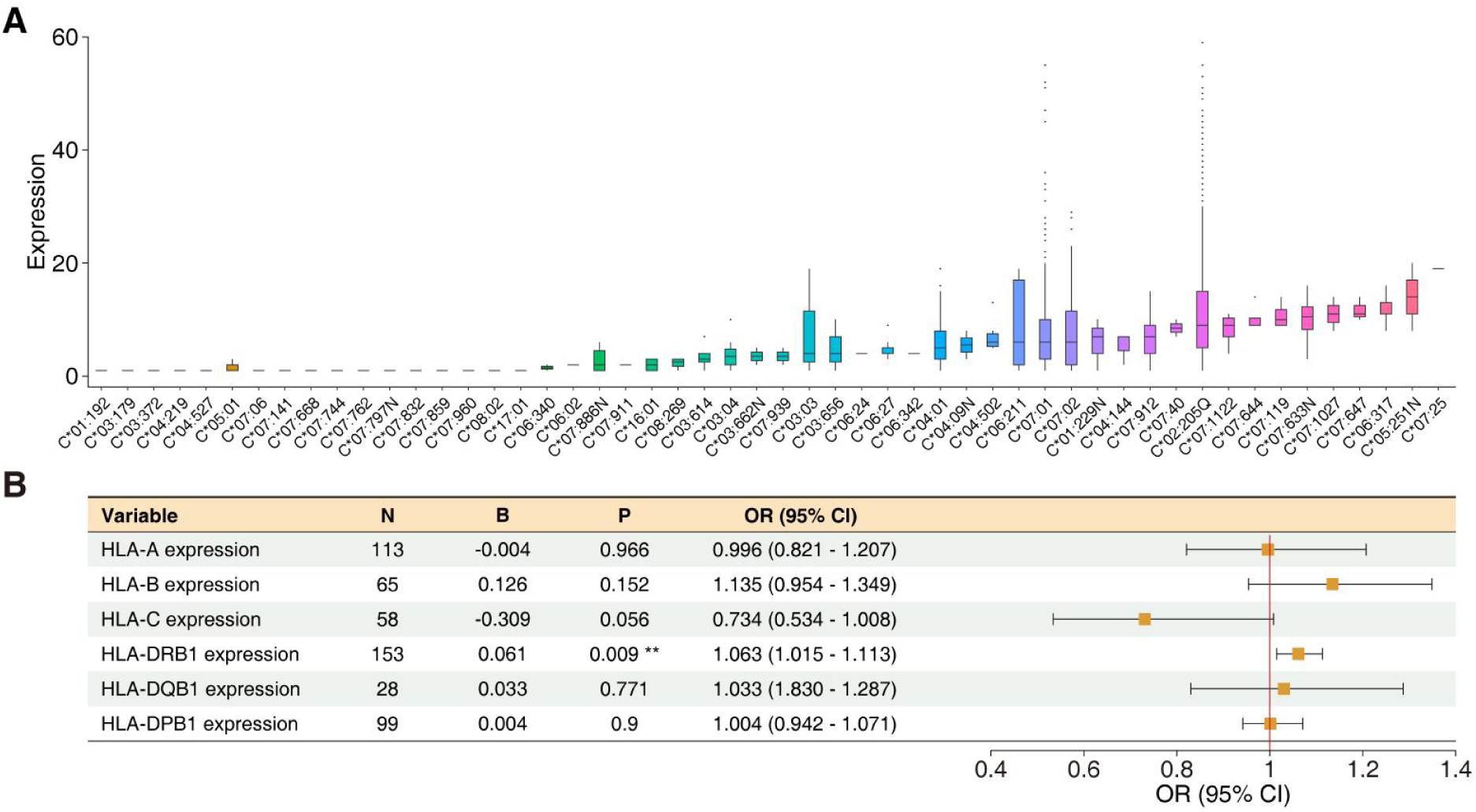
Correlation of HLA allele expression in cDCs with aGVHD development in HLA-matched patients. (**A**) Expression levels of HLA-C alleles in peripheral blood cDCs. (**B**) Correlation between HLA gene expression levels and aGVHD onset in HLA-matched patients. N, number of patients; B, regression coefficient; OR, odds ratio; CI, confidence interval. \*\**P* < 0.01.

To exclude confounding effects from donor-recipient HLA disparity, we restricted the analysis to 226 HLA-matched transplant cases. For each HLA gene, we screened the individuals with available expression data for the corresponding genotype and performed logistic regression to analyze the association between the gene expression level and post-transplant aGVHD occurrence. Among all six HLA genes, only *HLA-DRB1* expression in cDCs showed a significant positive correlation with aGVHD risk (OR = 1.063, *P* = 0.009) (**Figure 4B**). Therefore, we used recipient *HLA-DRB1* expression as a surrogate marker for the capacity to present miHAs in subsequent analyses.

### Combining DRs_HED with HLA allele expression improved the prediction of aGVHD onset

The activation of donor-specific T-cell receptors (TCRs) during aGVHD depends not only on HLA divergence but also on the expression levels of recipient HLA molecules on APCs. Previous studies have demonstrated that expression levels of mismatched *HLA-C* and *HLA-DPB1* influence aGVHD occurrence^32,34^. Therefore, integrating DRs_HED with allele-specific HLA expression is expected to more accurately reflect the degree of donor T-cell activation.

In our logistic regression analysis of the association between single-gene DRs_HED and aGVHD occurrence, we incorporated the expression level of the corresponding recipient-specific allele in cDCs as an additional variable. Since both variables may jointly influence disease pathogenesis, we further included their interaction term into the model. Additionally, we added the recipient’s *HLA-DRB1* expression level to account for the role of miHAs in aGVHD, thereby enabling a more comprehensive assessment of factors influencing aGVHD. Ultimately, the logistic regression model incorporated four variables: single-gene DRs_HED, the corresponding recipient-specific allele expression level, their interaction term, and recipient *HLA-DRB1* expression. This approach allowed us to evaluate the combined effect of DRs_HED and HLA expression on aGVHD development and to compare this model against models based solely on DRs_HED, assessing whether integrating both HLA disparity and expression improves predictive performance.

For DRs_HED, the DRs_HED of *HLA-A* (*P* = 0.003), *-B* (*P* = 0.002), *-C* (*P* = 0.001), *-DRB1* (*P* = 0.017), and *-DQB1* (*P* = 0.032) were all significantly associated with aGVHD occurrence, with the model area under the curve (AUC) values of 0.559, 0.566, 0.559, 0.543, and 0.543, respectively, indicating that the model considering only DRs_HED had some predictive ability for aGVHD (**Table 2**). However, when the model was expanded to include the additional 3 variables mentioned above, the AUC improved markedly from 0.551 ± 0.012 to 0.607 ± 0.012. These results demonstrate that incorporation of HLA expression data significantly enhances the model’s predictive performance.

**Table 2.**
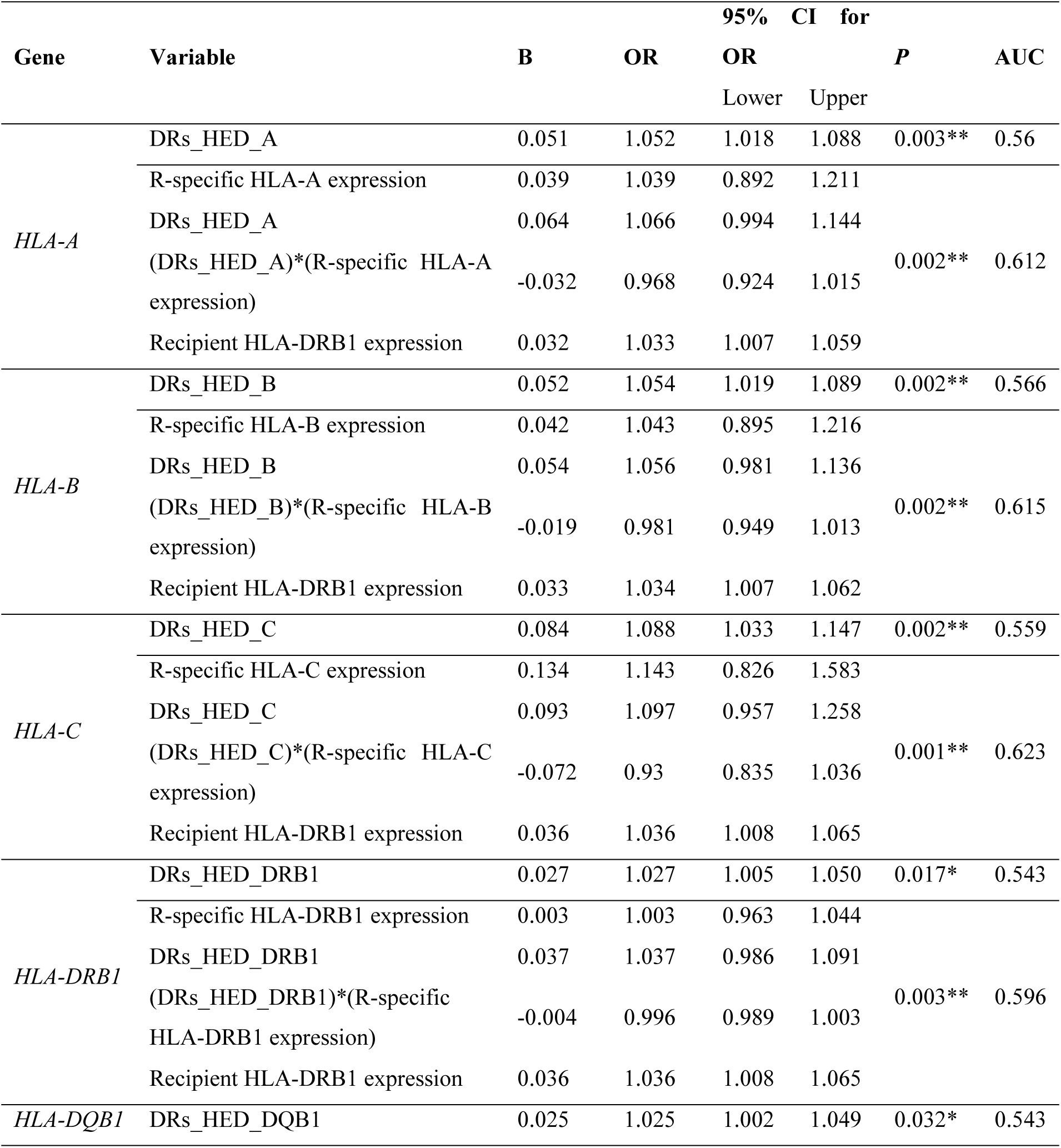
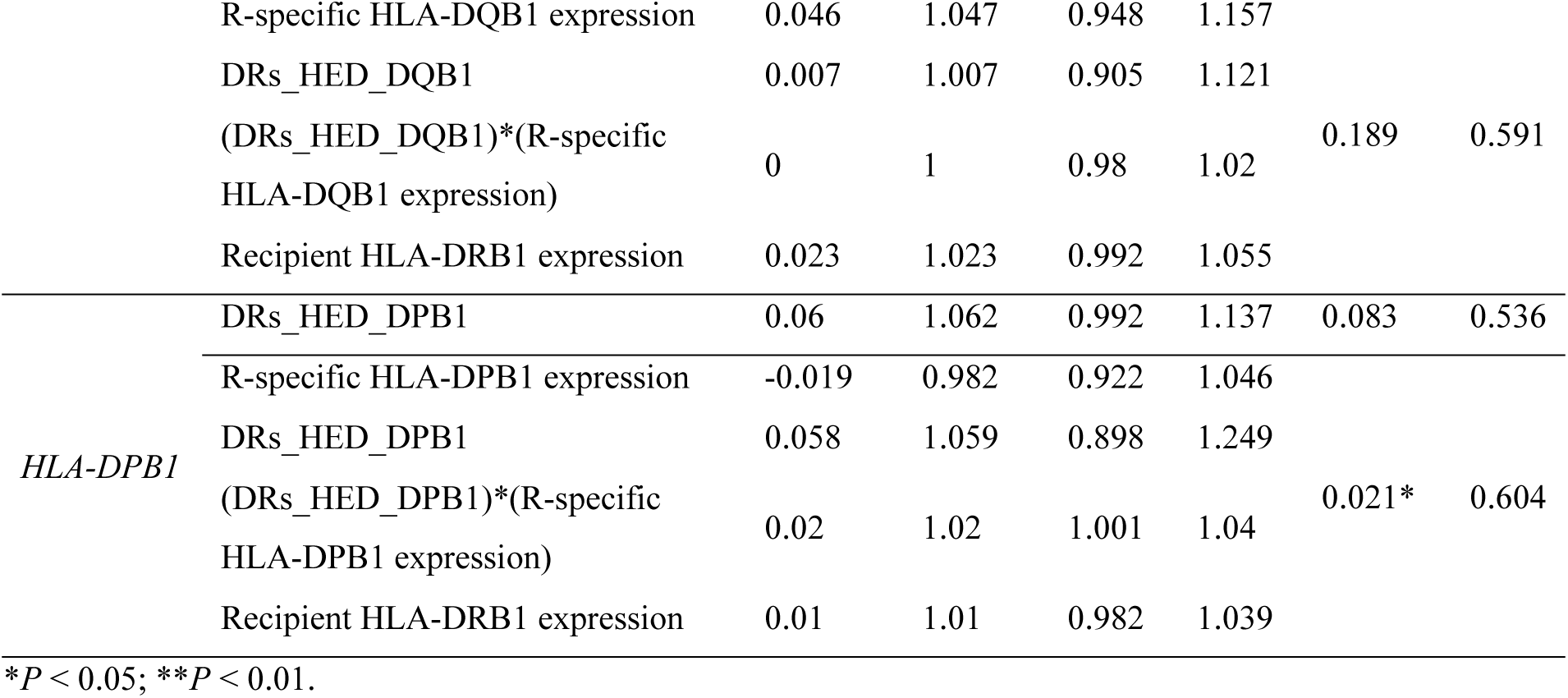
Predictive performance of base (DRs_HED only) and extended (DRs_HED + expression) models for aGVHD.

## DISCUSSION

Accurate prediction of aGVHD after aHSCT is crucial for optimal donor selection and effective prevention strategies. However, as haploidentical aHSCT becomes increasingly common, an effective prediction system is still lacking. Building on a refined quantification of donor-recipient HLA divergence, our retrospective cohort study of 774 patients demonstrates that both DRs_HED and HLA gene expression significantly influence the risk, organ specificity, and severity of aGVHD after aHSCT.

Donor-recipient HLA disparity is a key risk factor for aGVHD. To overcome limitations of earlier HED calculations for aGVHD prediction, we established the DRs_HED algorithm, which computes the average HED between the recipient-specific allele and both donor alleles, reflecting how recipient APCs activate donor T cells. Compared with earlier methods, DRs_HED resolved inconsistencies observed in prior studies and demonstrated higher predictive accuracy for aGVHD^30,39^. Furthermore, the independent contributions of multiple HLA loci underscore the polygenic nature of alloreactivity, a concept often underestimated in frequency-based HLA matching studies.

The impact of DRs_HED on aGVHD exhibited both locus universality and organ specificity. DRs_HED of all HLA class I genes and some class II genes (*HLA-DRB1* and *-DQB1*) was significantly associated with aGVHD risk and severity, with *HLA-C* having the strongest effect. DRs_HED of *HLA-DPB1* was not significantly correlated, possibly because some early patients lacked *HLA-DPB1* typing. Particularly noteworthy is that the association between DRs_HED and skin aGVHD was the most extensive and robust, with some association also existing with liver aGVHD, but no significant association with gastrointestinal aGVHD. This organ-specific pattern may stem from variations in HLA expression on primarily affected epithelial cells and vascular endothelial cells, local immune microenvironments, or organ-specific T cell homing mechanisms, warranting further study^40,41^.

Prior studies linked high HLA-C and HLA-DPB1 expression to aGVHD risk, but often used lymphoblastoid cell lines or T cells for expression detection and used antibodies cross-reactive with HLA-A and B to detect HLA-C, which complicates result interpretation^32,34,42^. In contrast, we leveraged allele-specific HLA expression data from cDCs at single-cell resolution, offering a biologically relevant model of post-transplant antigen presentation and a clearer understanding of HLA expression effects on aGVHD.

This study not only established the HED algorithm in the context of aGVHD but also combined it with HLA expression in cDCs, which play a central role in the initiation of aGVHD, improving the aGVHD prediction. Although most HLA loci (except *HLA-DRB1*) showed no significant association with aGVHD risk in fully matched transplants, in haploidentical settings, there was a significant synergistic effect between high recipient mismatched allele expression and high DRs_HED. Incorporation *HLA-DRB1* expression as a proxy for miHAs presentation by recipient APCs allowed a more comprehensive model which improved predictive performance for aGVHD with a mean AUC of 0.607 across HLA loci. Although its overall discriminative ability remains moderate, the stable statistical association, even without including clinical variables such as age and sex, supports DRs_HED as a core indicator of alloimmune responses.

Several limitations should be noted. First, the retrospective design may involve selection and information bias. Second, HLA expression data were derived from healthy donor cDCs rather than patient samples, and not all genotypes of studied patients were covered, restricting integrative analysis of multiple loci and predictive performance of the model. Future studies should establish inflammatory-condition expression databases of HLA allele in cDCs. In addition, recipients’ 6/8-digit HLA typing will facilitate more accurate determination of HLA allele expression levels and better prediction performance. At last, mRNA levels of HLA alleles as a surrogate for their cell surface protein levels needs to be confirmed in the future.

In summary, we present a refined measure of donor-recipient HLA disparity in the context of aGVHD and show that combining it with cDC-based allele-specific expression improves aGVHD risk prediction. These findings provide a new important perspective for understanding the immunobiological mechanisms of aGVHD, and also offers a basis for future development of precision donor selection strategies and individualized aGVHD risk stratification systems.

## METHODS

### Study design and study participants

This retrospective cohort study included 774 patients who underwent aHSCT between January 2013 and December 2024 at the First Affiliated Hospital of Xi’an Jiaotong University and the First Affiliated Hospital of Nanchang University. Inclusion criteria comprised the availability of complete high-resolution (4-digit) HLA typing for both donors and recipients, and comprehensive clinical follow-up data. Exclusion criteria included incomplete HLA data, non-aHSCT, or lack of post-transplant follow-up data. This study was approved by the Ethics Committee of the First Affiliated Hospital of Xi’an Jiaotong University and the First Affiliated Hospital of Nanchang University, in accordance with the Declaration of Helsinki. Written informed consent was obtained from all participants or their legal guardians.

### Data collection and variable definitions

The collected clinical variables, all extracted from the electronic medical record system, included recipient age, sex, diagnosis, pre-transplant disease remission status (complete remission (CR) or minimal residual disease (MRD)), conditioning regimen, donor age and sex, stem cell source, donor–recipient relationship, HLA matching degree, and donor-recipient cytomegalovirus (CMV) serostatus. Outcome recorded during follow-up comprised relapse, overall survival (OS), and relapse-free survival (RFS). aGVHD was diagnosed and graded (I–IV) within 100 days post-transplantation according to the Glucksberg criteria^43^. Organ-specific aGVHD (skin, gastrointestinal tract, and liver) was confirmed by clinicians based on clinical manifestations and histological evaluation.

### Calculation of Differential HLA Evolutionary Divergence (HED)

The HED score was calculated as the Grantham distance between the peptide-binding domains (exons 2 and 3 for *HLA-A*, *-B*, and *-C*; exon 2 for *HLA-DRB1*, *-DQB1*, and *-DPB1*) of the two alleles at each HLA locus. HED is calculated using the equation (2) as follows:

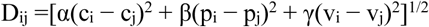

Here, i and j represent paired amino acids at the same position in the two allele sequences, c, p, and v represent composition, polarity, and molecular volume, respectively, and α, β, and γ denote their corresponding weighting coefficients. Recipient HED and donor-recipient HED were calculated as previously described^21,26,28^. Briefly, Recipient HED calculates the HED between the two alleles of the recipient at the same locus, while donor-recipient HED calculates the sum of HED between the recipient’s two alleles and the donor’s two alleles at the same locus. For all HED classification, the median HED score served as the threshold to define high and low HED groups.

### Expression of HLA alleles in cDC

The OneK1K consortium provides a population-scale single-cell RNA-seq dataset from Tasmania, Australia, comprising 982 individuals^44^. For each participant, an average of about 1,300 peripheral blood mononuclear cells (PBMCs) were profiled. After sequencing, the data were processed with rigorous quality control, normalization, and variance stabilization using sctransform^45^. Cell types were then annotated into 14 categories with scPred^46^, as previously described^44^. Building on this resource, we evaluated HLA allele expression in two sequential steps: first determining HLA alleles using ArcasHLA^47^, then quantifying allele-specific expression in cDCs at single-cell resolution using scHLAcount^48^.

### Statistical analysis

Pearson correlation analysis was used to assess correlations between DRs_HED values across HLA loci. Univariate binary logistic regression was employed to evaluate association of HED values with the risk of aGVHD, organ-specific aGVHD, and relapse. Ordinal logistic regression was applied to assess associations with aGVHD severity (grades I-IV). Cumulative incidence of aGVHD was estimated using the Kaplan-Meier method, with significance assessed by log-rank tests. Overall survival (OS) and relapse-free survival (RFS) probabilities were also calculated by Kaplan-Meier analysis. Univariate Cox regression was used to assess the hazard ratio (HR) for each variable. All statistical analyses were performed using R version 4.5.1 with a two-sided significance level set at 0.05.

## Acknowledgements

NA.

## Ethics Approval

This study was approved by the Ethics Committee of the First Affiliated Hospital of Xi’an Jiaotong University and the First Affiliated Hospital of Nanchang University, in accordance with the Declaration of Helsinki.

## Data Availability

The original contributions presented in the study are included in the article/supplementary material. Further inquiries can be directed to the corresponding author/s.

## Code Availability

This study did not generate new unique code.

## Author contributions

Conceptualization: Y.C., P.G.

Methodology: Y.C., P.G.

Software: H.G., Y.L., J.Q.

Validation: X.W., P.G.

Formal Analysis: Y.C., H.G.

Investigation: All authors.

Resources: P.H., X.W., F.L.

Data Curation: W.Z., Y.Z., Y.H., Y.L., S.Y., Y.G., X.W.

Writing: Y.C., P.G.

Visualization: Y.C., Z.L., P.G.

Supervision: P.H., X.W., F.L., P.G.

Project Administration: Y.C., P.G.

Funding Acquisition: P.G.

---

**Figure S1.**
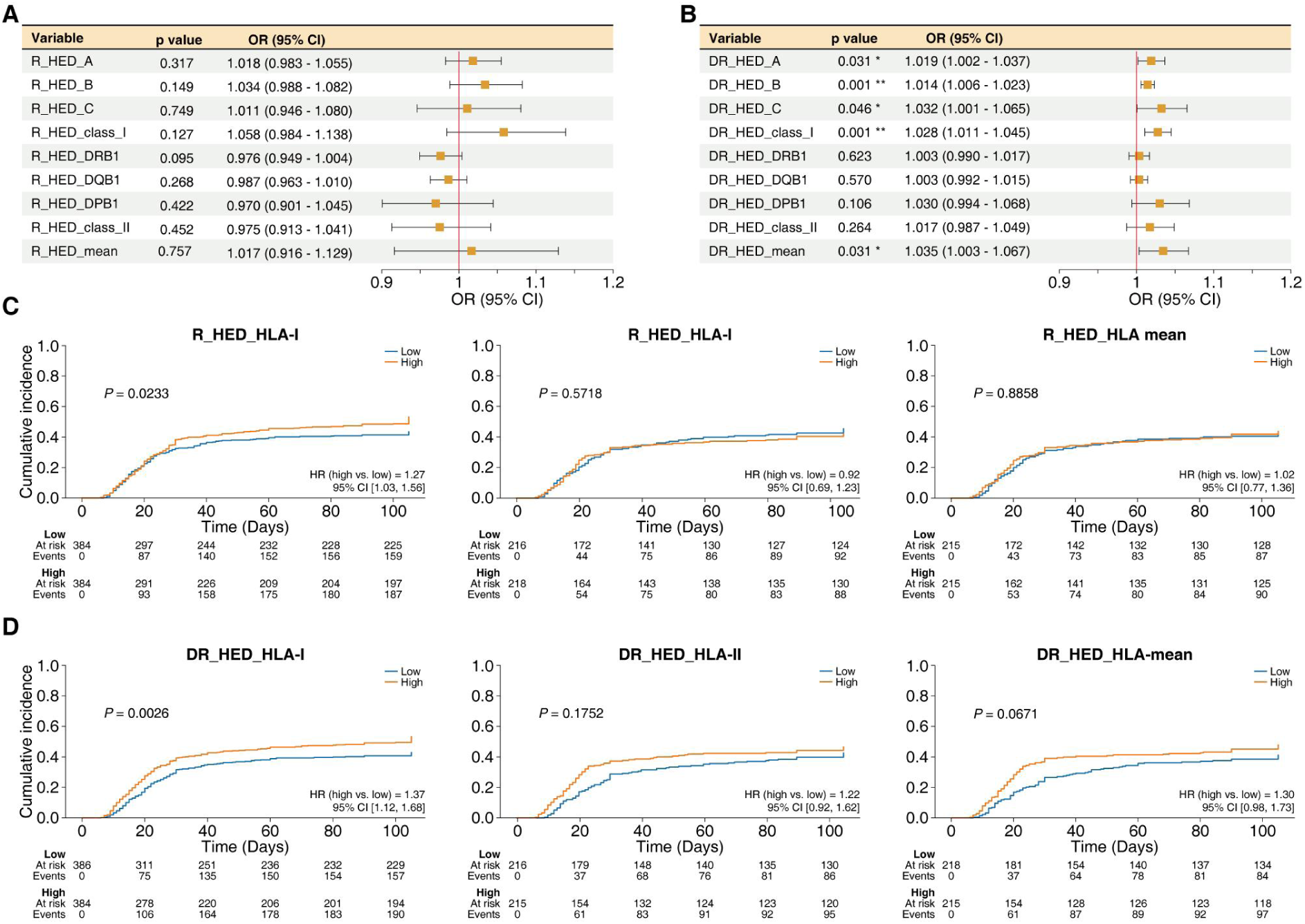
Association of R_HED and DR_HED with aGVHD onset. (A, B) Binary logistic regression analysis of the association between aGVHD occurrence and individual or averaged R_HED (A) or DR_HED (B) across six HLA loci. (C&D) Comparison of aGVHD cumulative incidence between high and low groups based on R_HED (C) or DR_HED (D) at HLA-I, -II, and average levels. \**P* < 0.05; \*\**P* < 0.01.

**Figure S2.**
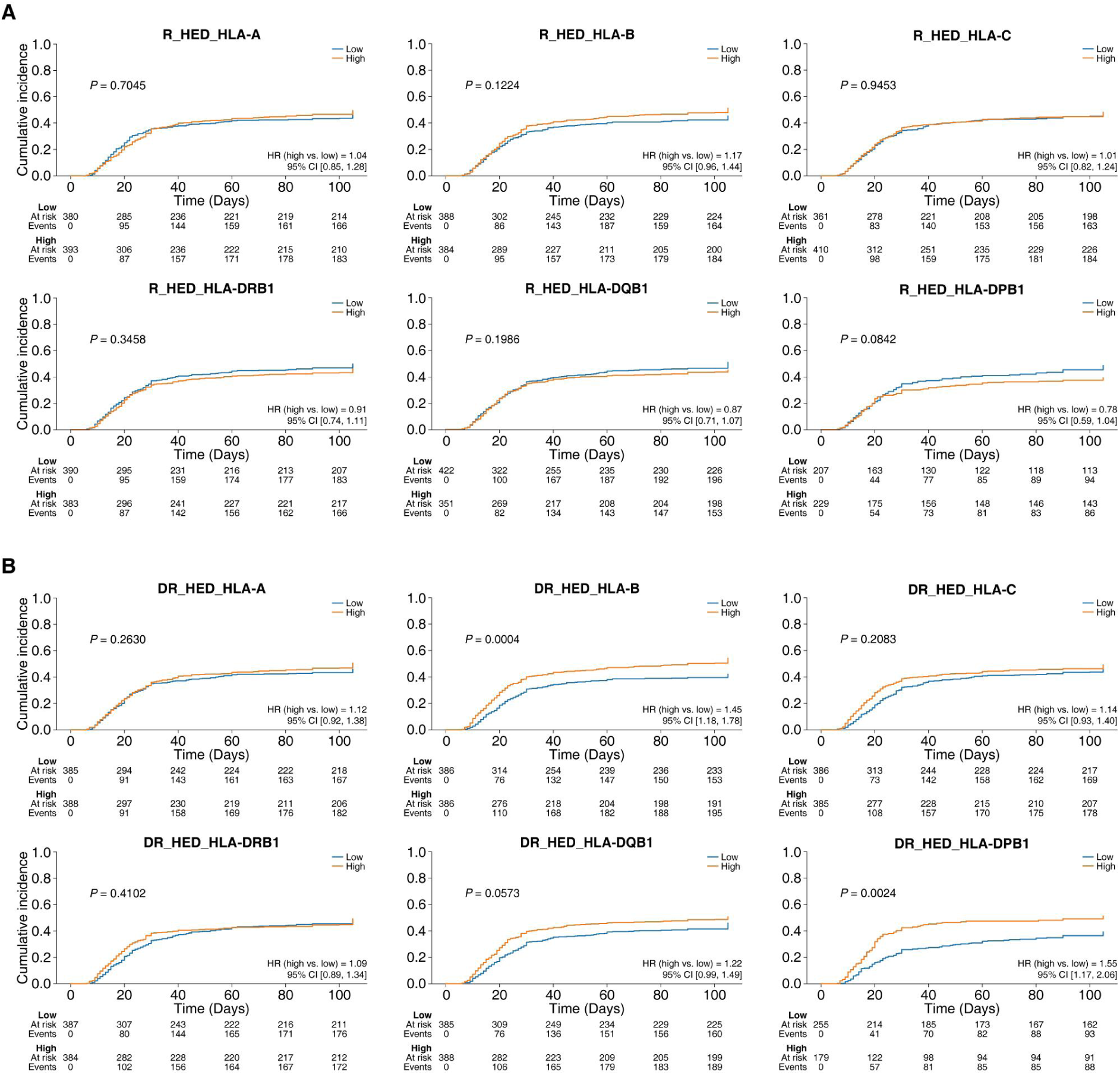
Comparison of aGVHD cumulative incidence between high and low groups based on R_HED (A) or DR_HED (B) of each individual HLA gene.

**Figure S3.**
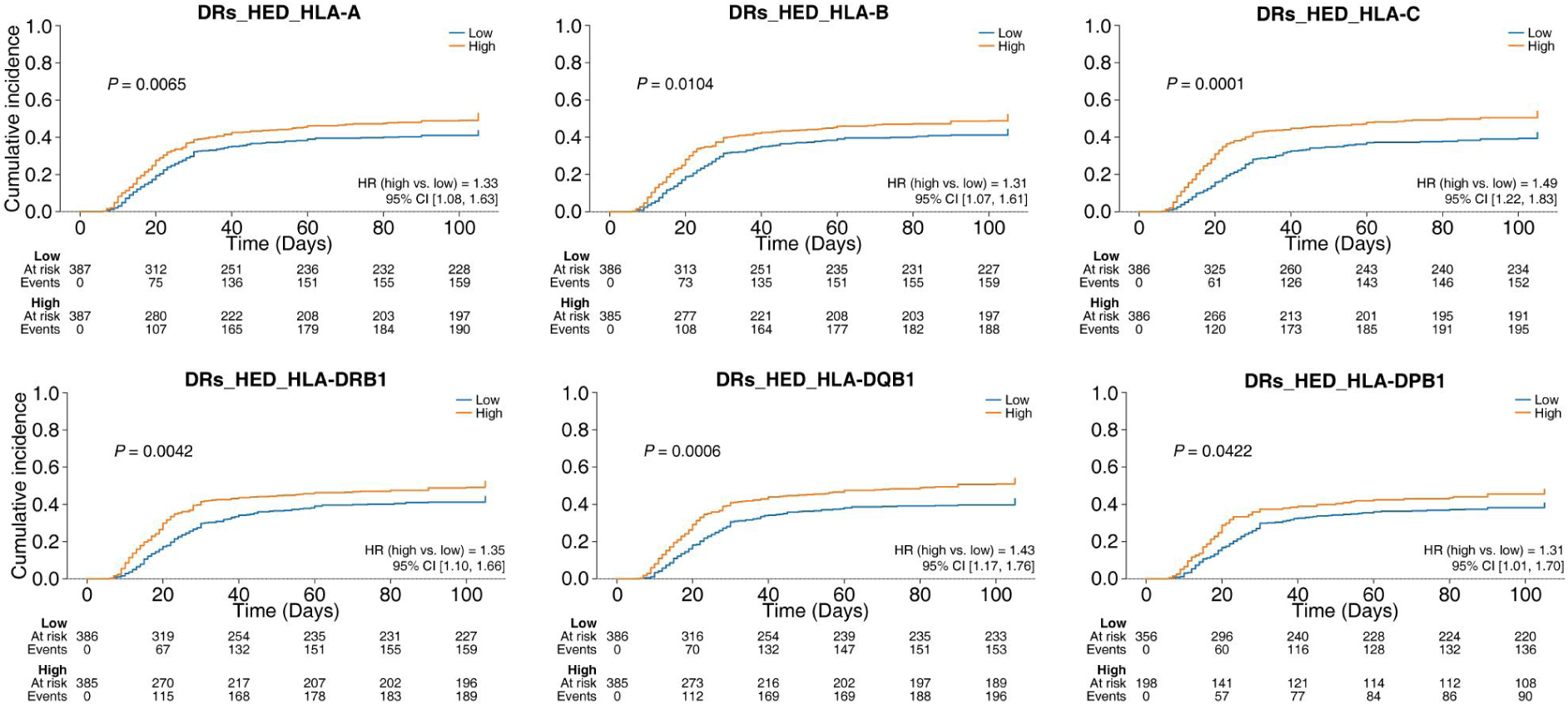
Comparison of cumulative aGVHD incidence between patient groups stratified by DRs_HED of each individual HLA gene.

**Figure S4.**
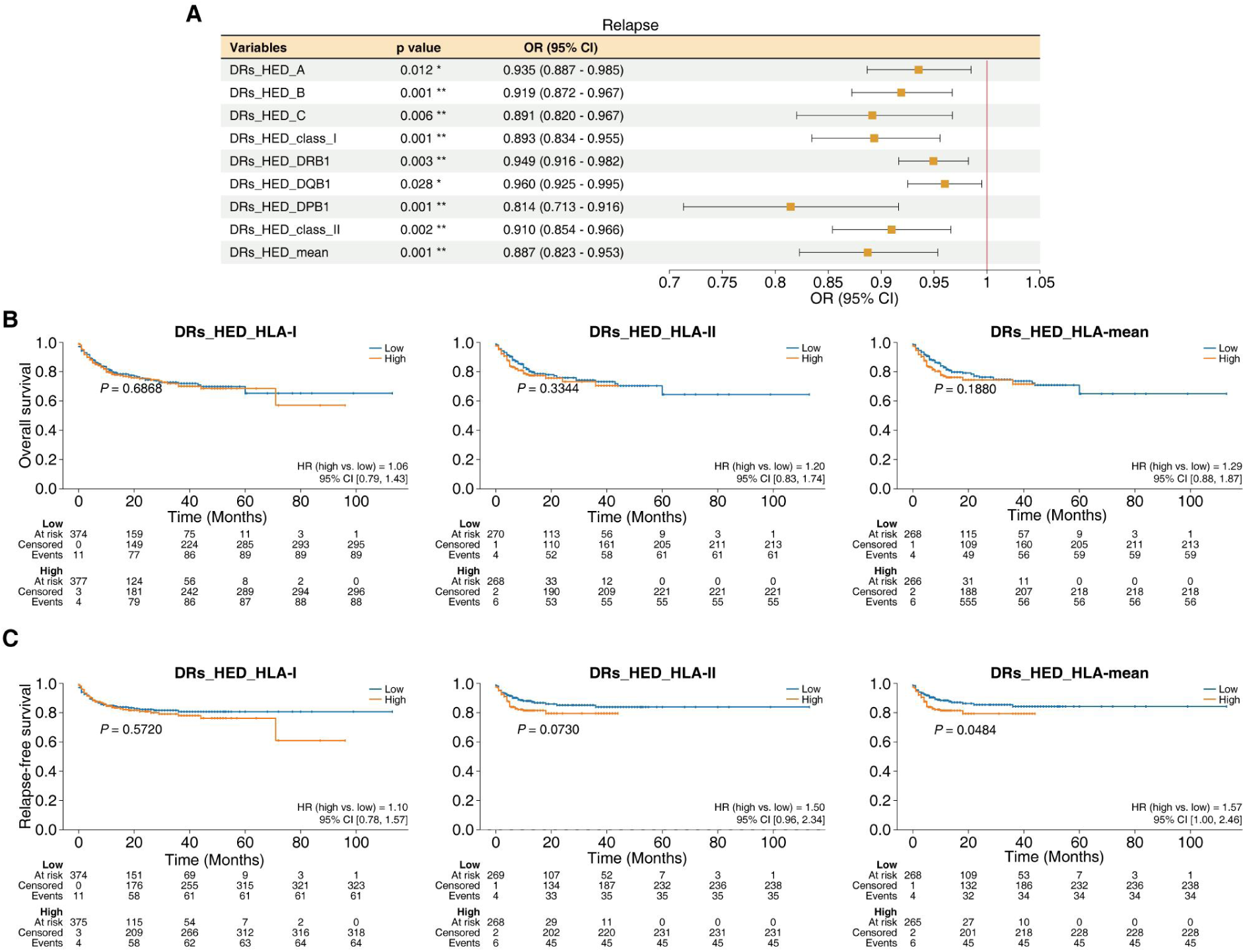
Correlation analysis between DRs_HED and clinical outcomes: relapse, overall survival and relapse-free survival. (**A**) Binary logistic regression analysis of the association between individual or average DRs_HED of 6 HLA genes and the relapse of primary disease. (**B&C**) Comparison of overall survival (B) and relapse-free survival (**C**) between patient groups stratified by average DRs_HED of HLA-I, HLA-II, and all 6 HLA genes.\**P* < 0.05; \*\**P* < 0.01.

## REFERENCES

1. Cieri, N., Maurer, K. & Wu, C. J. 60 Years Young: The Evolving Role of Allogeneic Hematopoietic Stem Cell Transplantation in Cancer Immunotherapy. Cancer Res 81, 4373–4384 (2021).

2. Zhang, X.-H. et al. The consensus from The Chinese Society of Hematology on indications, conditioning regimens and donor selection for allogeneic hematopoietic stem cell transplantation: 2021 update. J Hematol Oncol 14, 145 (2021).

3. Zeiser, R. & Blazar, B. R. Acute Graft-versus-Host Disease - Biologic Process, Prevention, and Therapy. N Engl J Med 377, 2167–2179 (2017).

4. Chen, Y.-B. et al. Vedolizumab for the prevention of intestinal acute GVHD after allogeneic hematopoietic stem cell transplantation: a randomized phase 3 trial. Nat Med 30, 2277–2287 (2024).

5. Hamilton, B. K. Current approaches to prevent and treat GVHD after allogeneic stem cell transplantation. Hematology Am Soc Hematol Educ Program 2018, 228–235 (2018).

6. Hill, G. R., Betts, B. C., Tkachev, V., Kean, L. S. & Blazar, B. R. Current Concepts and Advances in Graft-Versus-Host Disease Immunology. Annu Rev Immunol 39, 19–49 (2021).

7. Malard, F., Holler, E., Sandmaier, B. M., Huang, H. & Mohty, M. Acute graft-versus-host disease. Nat Rev Dis Primers 9, 27 (2023).

8. Biavasco, F. et al. Therapy response of glucocorticoid-refractory acute GVHD of the lower intestinal tract. Bone Marrow Transplant 57, 1500–1506 (2022).

9. Mo, X., Pei, X. & Huang, X. Optimization of T-cell-replete haploidentical hematopoietic stem cell transplantation: the Chinese experience. Haematologica 110, 562–575 (2025).

10. Guo, H. et al. CD8+HLA-DR+CD27+ T cells define a population of naturally occurring regulatory precursors in humans. Sci Adv 11, eadw1702 (2025).

11. Tamaki, M. et al. Associations between acute and chronic graft-versus-host disease. Blood Adv 8, 4250–4261 (2024).

12. Aharon, A. et al. MIF functional polymorphisms are associated with acute GVHD progression and steroid-refractoriness. Front Immunol 16, 1504976 (2025).

13. Wang, J., Liu, Y., Zhu, H. & Miao, K. Impact of HLA locus mismatch on peripheral blood allogeneic hematopoietic stem cell transplantation from unrelated donors using an ATG-based GVHD prophylaxis strategy. Blood Res 60, 34 (2025).

14. Arrieta-Bolaños, E. et al. Human Leukocyte Antigen Mismatching and Survival in Contemporary Hematopoietic Cell Transplantation for Hematologic Malignancies. J Clin Oncol 42, 3287–3299 (2024).

15. Iwasaki, M. et al. Impact of HLA Epitope Matching on Outcomes After Unrelated Bone Marrow Transplantation. Front Immunol 13, 811733 (2022).

16. Koyama, M. & Hill, G. R. Mouse Models of Antigen Presentation in Hematopoietic Stem Cell Transplantation. Front Immunol 12, 715893 (2021).

17. Kawase, T. et al. Identification of human minor histocompatibility antigens based on genetic association with highly parallel genotyping of pooled DNA. Blood 111, 3286–3294 (2008).

18. Spierings, E. Minor histocompatibility antigens: past, present, and future. Tissue Antigens 84, 374–360 (2014).

19. Likasitwatanakul, P. et al. HLA class I expression shapes the tumor immune microenvironment and influences prognosis in prostate cancer. Prostate Cancer Prostatic Dis 10.1038/s41391-025-01045-9 (2025) doi:10.1038/s41391-025-01045-9.

20. Viard, M. et al. Impact of HLA class I functional divergence on HIV control. Science 383, 319–325 (2024).

21. Chowell, D. et al. Evolutionary divergence of HLA class I genotype impacts efficacy of cancer immunotherapy. Nat Med 25, 1715–1720 (2019).

22. Pierini, F. & Lenz, T. L. Divergent Allele Advantage at Human MHC Genes: Signatures of Past and Ongoing Selection. Mol Biol Evol 35, 2145–2158 (2018).

23. Lu, Z. et al. Germline HLA-B evolutionary divergence influences the efficacy of immune checkpoint blockade therapy in gastrointestinal cancer. Genome Med 13, 175 (2021).

24. Féray, C. et al. Donor HLA Class 1 Evolutionary Divergence Is a Major Predictor of Liver Allograft Rejection : A Retrospective Cohort Study. Ann Intern Med 174, 1385–1394 (2021).

25. Mazzola, A. et al. HLA evolutionary divergence effect on bacterial infection risk in cirrhotic liver transplant candidates. Clin Immunol 270, 110399 (2025).

26. Roerden, M. et al. HLA Evolutionary Divergence as a Prognostic Marker for AML Patients Undergoing Allogeneic Stem Cell Transplantation. Cancers (Basel) 12, 1835 (2020).

27. Daull, A.-M. et al. Class I/Class II HLA Evolutionary Divergence Ratio Is an Independent Marker Associated With Disease-Free and Overall Survival After Allogeneic Hematopoietic Stem Cell Transplantation for Acute Myeloid Leukemia. Front Immunol 13, 841470 (2022).

28. Cao, X.-Y., Zhou, H.-F., Liu, X.-J. & Li, X.-B. Human leukocyte antigen evolutionary divergence as a novel risk factor for donor selection in acute lymphoblastic leukemia patients undergoing haploidentical hematopoietic stem cell transplantation. Front Immunol 15, 1440911 (2024).

29. Solh, M. et al. HLA evolutionary divergence (HED) informs the effect of HLA-B mismatch on outcomes after haploidentical transplantation. Bone Marrow Transplant 59, 1433–1439 (2024).

30. Merli, P. et al. Human leukocyte antigen evolutionary divergence influences outcomes of paediatric patients and young adults affected by malignant disorders given allogeneic haematopoietic stem cell transplantation from unrelated donors. Br J Haematol 200, 622–632 (2023).

31. Gong, X. & Karchin, R. Pan-Cancer HLA Gene-Mediated Tumor Immunogenicity and Immune Evasion. Mol Cancer Res 20, 1272–1283 (2022).

32. Petersdorf, E. W. et al. HLA-C expression levels define permissible mismatches in hematopoietic cell transplantation. Blood 124, 3996–4003 (2014).

33. Johansson, T., Partanen, J. & Saavalainen, P. HLA allele-specific expression: Methods, disease associations, and relevance in hematopoietic stem cell transplantation. Front Immunol 13, 1007425 (2022).

34. Petersdorf, E. W. et al. High HLA-DP Expression and Graft-versus-Host Disease. N Engl J Med 373, 599–609 (2015).

35. Petersdorf, E. W. et al. Role of HLA-DP Expression in Graft-Versus-Host Disease After Unrelated Donor Transplantation. J Clin Oncol 38, 2712–2718 (2020).

36. Fuchs, E. J. et al. Double unrelated umbilical cord blood vs HLA-haploidentical bone marrow transplantation: the BMT CTN 1101 trial. Blood 137, 420–428 (2021).

37. Zhang, Y., Louboutin, J.-P., Zhu, J., Rivera, A. J. & Emerson, S. G. Preterminal host dendritic cells in irradiated mice prime CD8+ T cell-mediated acute graft-versus-host disease. J Clin Invest 109, 1335–1344 (2002).

38. Lee, S. et al. Defining a TCF1-expressing progenitor allogeneic CD8+ T cell subset in acute graft-versus-host disease. Nat Commun 14, 5869 (2023).

39. Le Grand, S. et al. HLA evolutionary divergence score after donor lymphocyte infusion following allogeneic hematopoietic stem cell transplantation. Hemasphere 9, e70088 (2025).

40. Koyama, M. & Hill, G. R. Alloantigen presentation and graft-versus-host disease: fuel for the fire. Blood 127, 2963–2970 (2016).

41. Schreder, A. et al. Differential Effects of Gut-Homing Molecules CC Chemokine Receptor 9 and Integrin-β7 during Acute Graft-versus-Host Disease of the Liver. Biol Blood Marrow Transplant 21, 2069–2078 (2015).

42. Thomas, R. et al. HLA-C cell surface expression and control of HIV/AIDS correlate with a variant upstream of HLA-C. Nat Genet 41, 1290–1294 (2009).

43. Schoemans, H. M. et al. EBMT-NIH-CIBMTR Task Force position statement on standardized terminology & guidance for graft-versus-host disease assessment. Bone Marrow Transplant 53, 1401–1415 (2018).

44. Yazar, S. et al. Single-cell eQTL mapping identifies cell type-specific genetic control of autoimmune disease. Science 376, eabf3041 (2022).

45. Hofstetter, L. & Messerli, F. H. Hypothyroidism and hypertension: fact or myth? Lancet 391, 29–30 (2018).

46. Alquicira-Hernandez, J., Sathe, A., Ji, H. P., Nguyen, Q. & Powell, J. E. scPred: accurate supervised method for cell-type classification from single-cell RNA-seq data. Genome Biol 20, 264 (2019).

47. Orenbuch, R. et al. arcasHLA: high-resolution HLA typing from RNAseq. Bioinformatics 36, 33–40 (2020).

48. Darby, C. A., Stubbington, M. J. T., Marks, P. J., Martínez Barrio, Á. & Fiddes, I. T. scHLAcount: allele-specific HLA expression from single-cell gene expression data. Bioinformatics 36, 3905–3906 (2020).

